# Blood DNA Methylation Signature for Incident Dementia: Evidence from Longitudinal Cohorts

**DOI:** 10.1101/2024.11.03.24316667

**Authors:** Wei Zhang, Juan I. Young, Lissette Gomez, Michael A. Schmidt, David Lukacsovich, Brian W. Kunkle, Xi Chen, Eden R. Martin, Lily Wang

**Author notes:** Corresponding Address: Lily Wang, Soffer Clinical Research Ctr, University of Miami Miller School of Medicine 1120 NW 14th St, Miami, Florida 33136-2107. Data used in the preparation of this article were obtained from the Alzheimer’s Disease Neuroimaging Initiative (ADNI) database (http://adni.loni.usc.edu). As such, the ADNI investigators contributed to the design and implementation of ADNI and/or provided data but did not participate in the analysis or writing of this report. A complete listing of ADNI investigators is available at: http://adni.loni.usc.edu/wp-content/uploads/how_to_apply/ADNI_Acknowledgement_List.pdf.

## Abstract

**INTRODUCTION:** Distinguishing between molecular changes that precede dementia onset and those resulting from the disease is challenging with cross-sectional studies.

**METHODS:** We studied blood DNA methylation (DNAm) differences and incident dementia in two large longitudinal cohorts: the Offspring cohort of the Framingham Heart Study (FHS) and the Alzheimer’s Disease Neuroimaging Initiative (ADNI) study. We analyzed blood DNAm samples from over 1,000 cognitively unimpaired subjects.

**RESULTS:** Meta-analysis identified 44 CpGs and 44 differentially methylated regions consistently associated with time to dementia in both cohorts. Our integrative analysis identified early processes in dementia, such as immune responses and metabolic dysfunction. Furthermore, we developed a Methylation-based Risk Score, which successfully predicted future cognitive decline in an independent validation set, even after accounting for age, sex, APOE ε4, years of education, baseline diagnosis, and baseline MMSE score.

**DISCUSSION:** DNA methylation offers a promising source of biomarker for early detection of dementia.

## BACKGROUND

Alzheimer’s disease and related dementias (ADRD) are a major public health problem with a substantial economic burden^1^. ADRD currently affects 8.1 million to 10.8 Americans in the United States ^2^, and this number is projected to rise as the population ages. The escalating healthcare demands of ADRD underscore the critical need for effective prevention, early diagnosis, and management approaches.

Given the difficulty in halting neurodegenerative processes once they begin, it is imperative to develop biomarkers that could identify individuals at high risk for developing AD while they are still cognitively unimpaired (CU). Such biomarkers can facilitate personalized medicine and the implementation of preventive lifestyle interventions, potentially delaying the onset of dementia. Recent studies showed that delaying the onset of dementia by only one year in the 70-74-year-old group could reduce prevalence by more than 10%^3^.

DNA methylation (DNAm) is an epigenetic mechanism influenced by both genetics and environment. We and others have shown that DNAm is integrally involved in Alzheimer’s dementia (AD) ^4–11^. Moreover, several recent studies demonstrated DNAm differences could be detected in blood samples of AD subjects^12–17^. In particular, our recent analysis of two large clinical AD datasets (ADNI and AIBL) revealed a number of blood DNAm differences consistently associated with AD diagnosis in both cohorts^7^.

To develop biomarkers that can assess individual risk for dementia, it’s important to distinguish between DNAm changes that precede dementia onset and those that result from the disease. To date, most studies of DNAm in dementia have used a cross-sectional design^6,7, 16–18^ with only a few using the longitudinal design. Encouragingly, two recent longitudinal studies detected DNAm changes in the blood several years before the onset of dementia symptoms ^19–22^. However, these studies were limited by their small sample sizes.

Here we studied DNAm and incident dementia by meta-analyzing two large longitudinal datasets from the Framingham Heart Study (FHS) and the Alzheimer’s Disease Neuroimaging Initiative (ADNI) studies, with a total of more than 1000 samples from independent subjects, free of dementia or mild cognitive impairment (MCI) at blood sample collection. All samples included in this meta-analysis were measured using the same Infinium MethylationEPIC Beadchip platform, and each dataset was analyzed using a uniform analytical pipeline. We identified CpGs and differentially methylated regions (DMRs) consistently associated with incident dementia in both cohorts. Moreover, we also performed comparative analysis incorporating results from integrative analysis of DNAm in the blood with gene expression, genetic variants, and brain DNA methylation. These analyses, along with gene set enrichment analysis, highlighted DNAm differences associated with immune responses and metabolic dysfunction in dementia. In addition to corroborating findings from previous studies using cross-sectional design, our analysis also nominated a number of novel DNAm differences, emphasizing the importance of using a longitudinal design to identify DNAm differences with a temporal relationship to the disease. Importantly, we developed a Methylation Risk Score (MRS) for dementia using the FHS dataset and successfully validated it through out-of-sample testing on the ADNI dataset, demonstrating DNAm is a plausible source of predictive biomarkers for dementia.

## METHODS

### Study datasets for meta-analysis

The FHS is a community-based transgenerational study that investigates the development of cardiovascular disease in Framingham, Massachusetts^23^. In the FHS, the Offspring cohort included subjects from the second generation and their spouses. Blood samples were collected from the FHS Offspring cohort at Exam 9 (denoted FHS9 hereafter) which took place between 2011 – 2014. We included samples from 907 self-reported non-Hispanic White subjects who are free of dementia at Exam 9.

In the FHS, participants undergo Mini–Mental State Examinations (MMSE) at each exam cycle, and they completed a 45-minute neuropsychological test every 5-6 years since 1999. If participants are flagged for possible cognitive impairment based on these assessments, they are invited for additional, annual neurological and neuropsychological evaluations. If two consecutive annual evaluations show improvement, participants return to the regular follow-up schedule ^24^. Details of dementia surveillance in the FHS were previously described in Satizabal et al. (2016) ^24^. A dementia review panel assesses all potential cases, and the diagnosis of dementia is based on DSM-IV (Diagnostic and Statistical Manual of Mental Disorders, fourth edition) criteria. DNA methylation data and dementia ascertainment were obtained from the dbGap database (accessions: phs000974.v5.p4 and pht010750.v2.p14).

The ADNI is a longitudinal study designed to study the progression of AD ^25^. For our analysis, we selected the earliest visit with available DNAm data for each subject, and additionally required that they were cognitively normal at that time. This resulted in a dataset of 216 self-reported non-Hispanic White subjects ^26^. These subjects were followed approximately every 6 months, which provides valuable information on disease progression. DNA methylation data and the dementia status of the subjects were obtained from the ADNI study website (<u>adni.loni.usc.edu</u>).

The endpoint of this study is dementia onset. The follow-up period was from the time of blood sample collection for DNA methylation measurement to the time of dementia onset. Follow-up was censored at the time of loss to follow-up, non-dementia death, or the final day of study follow-up.

### Pre-processing of DNA methylation data

DNA methylation samples from both FHS9 and ADNI were measured using the same Illumina HumanMethylation EPIC v1 bead chips. Supplementary Table 1 shows the number of CpGs and samples at each quality control (QC) step. The FHS9 and ADNI datasets were pre-processed separately. For each dataset, the QC of probes involved several steps. First, we selected probes with a detection *P-*value < 0.01 in 90% or more of the samples. A small *P-*value indicates a significant difference between the signals in the probes and the background noise. Next, we selected probes that start with “cg”, and using the function rmSNPandCH from the DMRcate R package, we removed probes that are located on X and Y chromosomes, are cross-reactive ^27^, or located close to single nucleotide polymorphism (SNPs) (i.e., an SNP with minor allele frequency (MAF) ≥ 0.01 was present in the last five base pairs of the probe).

For QC of the samples, we first removed samples with bisulfite conversion rate lower than 85%, as well as samples for which the DNAm predicted sex status differed from the recorded sex status. The sex prediction was performed using the getSex function from the minfi R package. In addition, we performed principal component analysis (PCA) using the 50,000 most variable CpGs to identify outliers. Samples outside the range of ±3 standard deviations from the mean of PC1 and PC2 were excluded.

The quality-controlled data was next normalized using the dasen method, as implemented in the wateRmelon R package ^28^. Immune cell type proportions (B lymphocytes, natural killer cells, CD4+ T cells, CD8+ T cells, monocytes, neutrophils, and eosinophils) were estimated using the EpiDISH R package ^29^. As in previous blood-based DNAm studies ^6,7,30^, granulocyte proportions were computed as the sum of neutrophils and eosinophils proportions since both neutrophils and eosinophils are classified as granular leukocytes. To correct batch effects from methylation plates, we used the BEclear R package ^31^.

Supplementary Figures 1-2 show that the first principal component (PC1) of methylation beta values was not significantly associated with covariates including follow-up duration, age, sex, education and smoking history of the subjects in ADNI and FHS9 datasets, with the exception that PC1 was significantly higher in males in the FHS9 dataset. All subsequent analyses were adjusted for sex along with other potential confounding factors.

### Association of DNA methylation at individual CpGs with dementia

To evaluate the relationship between incident dementia and DNA methylation, we conducted Cox proportional regression analyses on both FHS and ADNI datasets separately, via the coxph function in the survival R package. For the FHS dataset, we used the model: Surv (follow-up time, status) ∼ methylation.beta + age + sex + immune cell-type proportions (B, NK, CD4T, Mono, Gran) where status indicates whether incident dementia occurred (1 = event occurred, 0 = censored) (Model 1). For the ADNI dataset, given the smaller sample size, we only included the first two principal components (PCs) of the immune cell-type proportions, which explained 90.0% variances in estimated immune cell-type proportions. Specifically, we fitted the model Surv (follow-up time, status) ∼ methylation.beta + age + sex + PC1 + PC2.

### Inflation assessment and correction

Genomic inflation factors (lambda values) were estimated using both the conventional approach ^32^ and the *bacon* method ^33^, which was proposed specifically for EWAS. For the FHS and ADNI datasets, the estimated bias was −0.009 and 0.041, respectively. For the estimated inflation, the lambda values (λ) using the conventional approach were 1.424 and 1.005, while the lambda values based on the bacon approach (λ.bacon) were 1.176 and 0.986 for the FHS and ADNI datasets, respectively (Supplementary Table 2).

We next applied genomic correction using the bacon method^33^, as implemented in the bacon R package, to obtain bacon-corrected effect sizes, standard errors, and *P-*values for each dataset. After bacon correction, the estimated bias were 4.78×10^-4^ and −6.18×10^-4^, and the estimated inflation factors were λ = 1.03 and 1.029, and λ.bacon = 1.01 and 1.00 for the FHS and ADNI datasets, respectively.

### Meta-analysis

To meta-analyze individual CpG results across both the FHS9 and ADNI datasets, we used the inverse-variance weighted fixed-effects model, implemented in the meta R package. As demonstrated by Rice et al. (2018), the fixed effects model can be interpreted as a weighted average of study-specific effects, regardless of whether the true study-specific effects are heterogeneous ^34^. The methylation beta values were rescaled into z-scores so that the estimated hazard ratios correspond to an increase in dementia risk associated with a one standard deviation increase in beta values. To correct for multiple comparisons, we computed the false discovery rate (FDR). We considered CpGs with an FDR less than 5% in meta-analysis of the FHS9 and ADNI datasets, with a consistent direction of change in estimated effect sizes, and a nominal *P-*value less than 0.05 in both datasets as statistically significant.

### Differentially methylated regions analysis

For region-based meta-analysis, we used the comb-p method ^35^. Briefly, comb-p takes single CpG *P*-values and locations of the CpG sites to scan the genome for regions enriched with a series of adjacent low *P*-values. In our analysis, we used *P*-values from the meta-analysis of the FHS9 and ADNI datasets as input for comb-p. We used parameter settings with --seed 0.05 and--dist 750 (a *P*-value of 0.05 is required to start a region and extend the region if another *P*-value was within 750 base pairs), which were shown to have optimal statistical properties in our previous comprehensive assessment of the comb-p software^36^. As comb-p uses the Sidak method to account for multiple comparisons, we selected DMRs with Sidak *P*-values less than 0.05. To help reduce false positives, we imposed two additional criteria in our final selection of DMRs: (1) the DMR also has a nominal *P*-value < 1×10^-5^; (2) all the CpGs within the DMR have a consistent direction of change in estimated effect sizes in the meta-analysis.

### Functional annotation of significant methylation associations

The significant methylation at individual CpGs and DMRs were annotated using both the Illumina (UCSC) gene annotation and Genomic Regions Enrichment of Annotations Tool (GREAT) software ^37^ which associates genomic regions with target genes. To assess the overlap between our significant CpGs and DMRs (CpG or DMR location +/-250bp) with enhancers, we used enhancer gene maps generated from 131 human cell types and tissues described in Nasser et al. (2021) ^38^. Specifically, we selected enhancer-gene pairs with “positive” predictions from the ABC model, which included only expressed target genes, did not include promoter elements, and had an ABC score higher than 0.015. In addition, we also required that the enhancer-gene pairs be identified in cell lines relevant to this study.

### Pathway analysis

To identify biological pathways enriched with significant DNA methylation differences, we used the methylRRA function in the methylGSA R package ^39^, which used *P*-values from the meta-analysis of FHS9 and ADNI datasets as input. Briefly, methylGSA first computes a gene-wise ρ value by aggregating *P*-values from multiple CpGs mapped to each gene. Next, the different number of CpGs on each gene is adjusted by Bonferroni correction. Finally, a Gene Set Enrichment Analysis ^40^ (in pre-rank analysis mode) is performed to identify pathways enriched with significant CpGs. We analyzed pathways in the KEGG and REACTOME databases. Pathways with FDR less than 0.05 were considered to be statistically significant.

### Integrative analyses with gene expression, genetic variants, and brain-to-blood correlations

To evaluate the effect of DNA methylation on the expression of nearby genes, we overlapped our dementia-associated CpGs, including both significant individual CpGs and those located within DMRs, with eQTm analysis results in Supplementary Tables 2 and 3 of Yao et al. (2021)^41^.

For correlation and overlap with genetic susceptibility loci, We searched for mQTLs in the blood using the GoDMC database^17^. To select significant blood mQTLs in GoDMC, we used the same criteria as the original study ^42^, that is, considering a cis *P-*value smaller than 10^-8^ and a trans *P-*value smaller than 10^-14^ as significant. The genome-wide summary statistics for genetic variants associated with dementia described in Bellenguez et al. (2022) ^43^ were obtained from the European Bioinformatics Institute GWAS Catalog under accession no. GCST90027158. Colocalization analysis was performed using the coloc R package.

To assess the correlation of dementia-associated CpGs and DMRs methylation levels in blood and brain samples, we used the London dataset, which consisted of 69 samples with matched PFC and blood samples^44^. We assessed the association of brain and blood methylation levels at dementia-associated CpGs using both an unadjusted correlation analysis with methylation beta values (*r*_*beta*_), and an adjusted correlation analysis using methylation residuals (*r*_*r*esid_), in which we removed the effect of estimated neuron proportions in brain samples (or estimated immune cell-type proportions in blood samples), array, age at death for brain samples (or age at blood draw for blood samples), and sex from DNA methylation *M*-values.

### Sensitivity analysis

In the first sensitivity analysis, we evaluated if dementia risk factors would likely confound the DNA methylation to dementia associations we observed. To this end, we first performed regression analysis to assess the association between dementia-associated CpGs (both significant individual CpGs and those located in DMRs) and dementia risk factors collected by the Framingham study, including diabetes, blood pressure, years of education, obesity, and smoking. Specifically, for each risk factor and each CpG, we fitted the model methylation.m.value ∼ risk factor + age + sex + cell type proportions (B, NK, CD4T, Mono, Gran). A risk factor is considered significantly associated with a CpG if its *P*-value is less than 0.05 in the above model (i.e., a significant risk factor for a CpG).

Next, the confounding effects of these significant risk factors for the dementia-associated CpGs were evaluated by fitting the Cox proportional regression model that expanded Model 1 above by additionally including the significant risk factor: Surv(incident dementia, follow-up time) ∼ methylation.beta + risk factor + age + sex + immune cell-type proportions (B, NK, CD4T, Mono, Gran).

In the second sensitivity analysis, to evaluate the impact of family structure in the discovery of significant CpGs, we computed the intraclass correlation coefficient (ICC) for dementia-associated CpGs, by fitting a random effects model methylation m-value ∼ random (family) to the FHS data for each CpG. The ICC was then estimated by 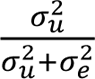 where σ^2^_u_ is estimated variance component for family random effects, and σ^2^ is the residual error. This analysis was implemented using the lmer function in the lme4 R package. We also compared the results of Model 1 with a model that accounts for family relationships in the Cox regression model using a kinship matrix. To this end, we first computed the kinship matrix using the R package kinship2 based on the pedigree information in the dbGap dataset (accession: pht000183.v13.p14). Next, for each dementia-associated CpGs, we fitted a mixed-effects Cox regression model with a random intercept, adjusting for the same covariates as in Model 1 above in the analysis of the FHS9 dataset. The variance-covariance matrix is determined using twice the value of the kinship matrix ^45^. The mixed-effects Cox models were implemented using the coxme R package.

The coMethDMR^46^ software was used to evaluate the robustness of genomic regions defined by the 44 DMRs identified by comb-p for their association with time to incident dementia. First, coMethDMR selected co-methylated sub-regions within for each region. We then summarized methylation beta values within these co-methylated sub-regions using medians and tested them against time to incident dementia. In the same way as in single CpG analyses, we adjusted for potential confounding factors, including age, sex, and blood cell-type composition. The dataset specific *P-*values for each genomic region were then combined across FHS9 and ADNI datasets using an inverse-variance weighted fixed effects meta-analysis model. DMRs with an FDR of less than 5% were considered significant.

In the last sensitivity analysis, to identify cases with dementia or MCI due to AD in the ADNI dataset, we use the variables DXMDUE and DXDUE in the DXSUM table. Similarly, we used the variable AD_STATUS in the FHS9 dataset to identify AD cases.

### Validation using independent datasets

To compare our results with previous findings, we searched dementia-associated CpGs (both significant individual CpGs and those located in DMRs) using the CpG Query tool in the MIAMI-AD database^47^. For input on phenotype, we selected “AD Biomarker”, “AD Neuropathology”, “Dementia Clinical Diagnosis”.

### Out-of-sample validation of Methylation Risk Score

First, we selected CpGs that achieved *P-*value < 10^-5^ using the model Surv (follow-up time, status) ∼ methylation.beta + age + sex + immune cell-type proportions (B, NK, CD4T, Mono, Gran) where status indicates whether incident dementia occurred (1 = event occurred, 0 = censored) in the FHS9 dataset. To estimate CpG weights, we applied ridge regression using the glmnet R package on the FHS9 dataset. The model parameters λ was optimized via five-fold cross validation based on Harrell C index. After tunning the model, the final model was fitted with λ = lambda.1se, i.e., the value of λ that gives the most regularized model such that the cross-validated C index is within one standard error of the maximum.

Next, we performed an out-of-sample validation using the ADNI dataset. For each subject at baseline, the MRS was computed by summing the methylation *M-*values for the 151 CpGs weighted by coefficients estimated from the ridge regression model described above. We then performed Cox regression analyses on the ADNI dataset to evaluate the association between baseline MRS and disease progression. This analysis was performed using the coxph function in the survival R package. Disease progression was defined as the conversion from CN to MCI or dementia, and from MCI to dementia. The model was adjusted for multiple covariates: Surv (Conversion event, follow-up time) ∼ MRS + age + sex + APOE ε4 status + years of education + baseline diagnosis + baseline MMSE score. Furthermore, we stratified the samples into four groups based on quartiles of their baseline MRS scores and visualized the conversion probabilities for the subjects in the first and last quartiles, after adjusting for covariate variables age, sex, APOE ε4 status, years of education, baseline diagnosis, and baseline MMSE score. The adjusted Kaplan-Meier curves were graphed using the ggadjustedcurves function from the survminer R package.

### Data and Code Availability

The genome-wide summary statistics have been deposited to the MIAMI-AD (DNA <u>M</u>ethylation in <u>A</u>ging and <u>M</u>ethylation in <u>AD</u>) database (https://miami-ad.org/). The scripts for the analyses performed in this study are available at https://github.com/TransBioInfoLab/blood-dnam-and-incident-dementia

## RESULTS

### Study cohorts

Our meta-analysis included a total of 1123 DNAm samples (measured using Illumina EPIC arrays, generated from blood samples of 907 Offspring cohort participants (496 females, 411 males) in the Framingham Heart Study (FHS) and 216 participants (108 females, 108 males) in the Alzheimer’s Disease Neuroimaging Initiative (ADNI) study (Table 1), who were free of dementia or MCI diagnosis at blood sample collection.

**Table 1:** Characteristics of subjects included in the meta-analysis of the Framingham Heart Study Exam 9 (FHS9) and ADNI cohorts. Dementia subjects are defined as those who developed dementia during the follow-up period; control subjects are those who did not develop dementia during the observation period or were lost to follow-up.

| Characteristics | Framingham Heart Study Offspring Cohort at Exam 9 |  |  |  | ADNI (Earliest visit with DNA methylation data) |  |  |  |
| --- | --- | --- | --- | --- | --- | --- | --- | --- |
|  | All<br>(n = 907) | Dementia<br>(n = 42) | Control<br>(n = 865) | P-value | All<br>(n = 216) | Dementia<br>(n = 18) | Control<br>(n = 198) | P-value |
| Follow up period in years |  |  |  |  |  |  |  |  |
| Av. (sd) | 4.98 (2.33) | 3.06 (1.84) | 5.07 (2.31) | $2.24 \times 10^{-8}$ | 5.97 (3.00) | 6.42 (2.72) | 5.92 (3.03) | 0.513 |
| Age, av. (sd) | 72.03 (8.18) | 79.26 (5.96) | 71.68 (8.12) | $2.77 \times 10^{-9}$ | 76.73 (6.62) | 74.92 (3.98) | 76.63 (6.75) | 0.368 |
| Sex, n (%) |  |  |  |  |  |  |  |  |
| Female | 496 (54.69) | 20 (47.62) | 476 (55.03) | 0.428 | 108 (50.00) | 10 (55.56) | 98 (49.50) | 0.806 |
| Male | 411 (45.31) | 22 (52.38) | 389 (44.07) |  | 108 (50.00) | 8 (44.44) | 100 (51.50) |  |
| Mean education year, av. (sd) | 14.38 (2.53) | 13.84 (2.39) | 14.41 (2.54) | 0.317 | 16.36 (2.67) | 16.11 (2.70) | 16.38 (2.67) | 0.522 |
| Smoking history, n (%) |  |  |  |  |  |  |  |  |
| Yes | 549 (60.60) | 29 (69.05) | 520 (60.19) | 0.332 | 92 (42.59) | 8 (44.44) | 84 (42.42) | 1.000 |

**Table 2.**
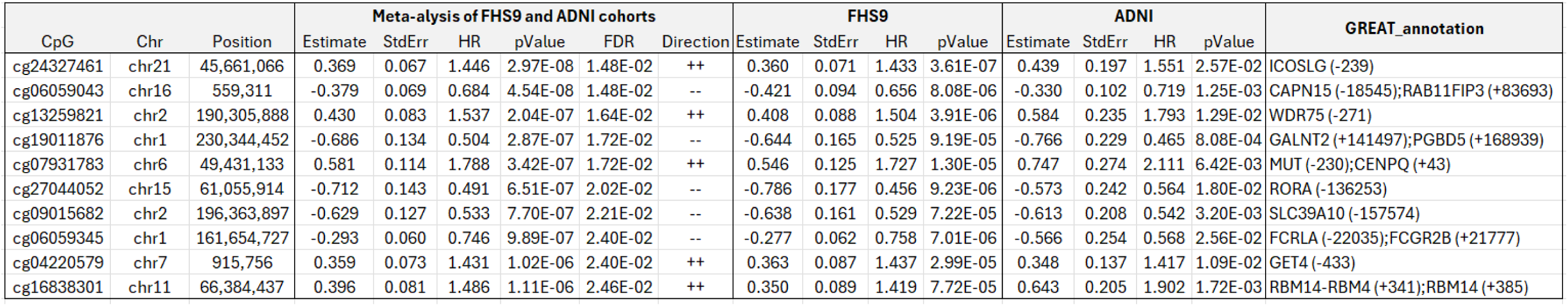
Top 10 most significant CpGs associated with incident dementia in the meta-analysis of blood samples in ADNI and Framingham Exam 9 (FHS9) datasets. Inverse-variance weighted fixed-effects meta-analysis models were used to combine cohort-specific results from Cox regression models that included covariate variables age, sex, and immune cell-type proportions. Hazard ratios (HR) describe changes in risk of dementia associated with a one standard deviation increase in methylation beta values after adjusting for covariate variables. We corrected batch effects using the BEclear R package, and corrected genomic inflation using the bacon R pakcage. Direction indicates hypermethylation (+) or hypomethylation (-) of the CpG associated with increased risk of dementia in FHS9 and ADNI datasets. In GREAT annotation, the numbers in parentheses indicate the distance from the TSS.

**Table 3.**
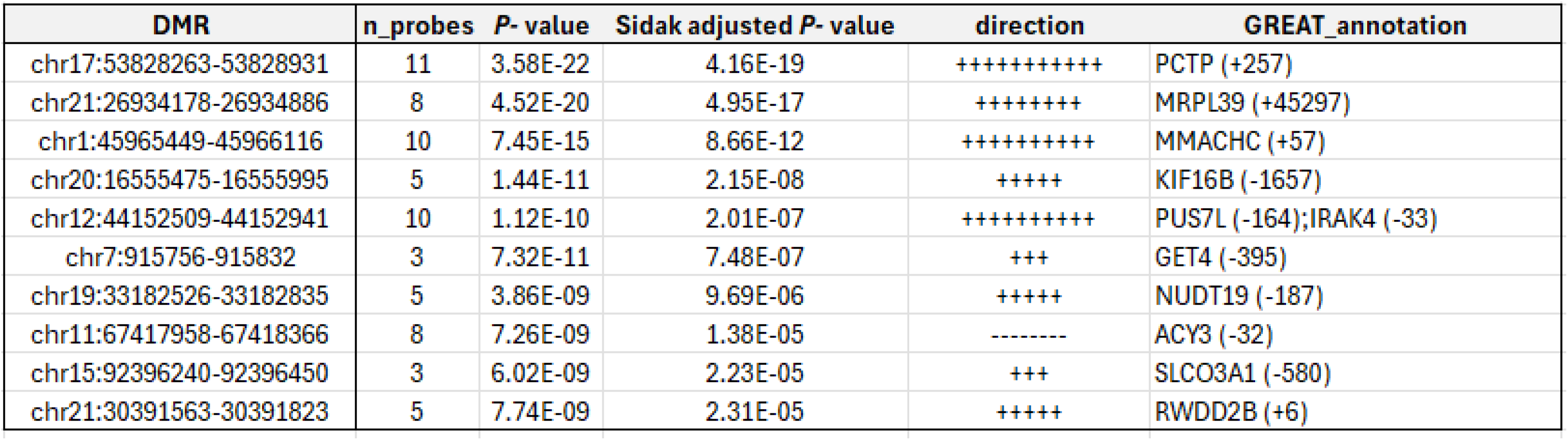
Top 10 most significant differentially methylated regions (DMRs) associated with incident dementia. Direction indicates directions of CpGs within each DMR in the meta-analysis of FHS9 and ADNI datasets, with hypermethylation (+) or hypomethylation (-) associated with increased risk of dementia. In GREAT annotation, the numbers in parentheses indicate the distance from the TSS.

In the FHS, the mean ages of the subjects at blood collection during Exam 9 was 72.03 ± 8.18 years. The subjects in FHS9 were followed up to 7.72 years after Exam 9, with an average follow-up of 4.98 ± 2.33 years, and 42 subjects developed dementia during this period. In particular, for those who did not develop dementia by the end of the observation period in 2018, the average follow-up duration was 5.07 ± 2.31 years (Table 1, Supplementary Figure 3).

In the ADNI, the mean age of the participants at the time of sample collection was 76.73 ± 6.62 years. These subjects were followed for up to 11.11 years, with an average follow-up of 5.97 ± 3.00 years, and 18 subjects developed dementia during this period. For those who did not develop dementia by the end of the observation period, the average follow-up duration was 5.92 ± 3.03 years (Table 1, Supplementary Figure 4). The percentages of non-smokers in FHS9 and ADNI were 39.40% and 57.41%, respectively. Subjects in both cohorts are highly educated, with an average of 14.38 years of education in FHS9 and 16.36 years in ADNI.

### Blood DNAm differences at individual CpGs and DMRs are significantly associated with incident dementia

After adjusting for age, sex, and immune cell type proportions, and correcting batch effects and genomic inflation (Methods), we identified 44 CpGs with a consistent direction of change in both FHS9 and ADNI datasets, a nominal *P-*value less than 0.05 in both datasets and an FDR < 0.05 in inverse-variance fixed-effects meta-analysis of the FHS9 and ADNI datasets (Figure 1, Supplementary Table 3, Supplementary Figure 5). These results remain robust in models that additionally included dementia risk factors and accounted for family structure in the FHS (see details below). For these 44 significant CpGs, the hazard ratios associated with one standard deviation change in methylation beta values ranged from 0.456 to 3.948 in the FHS9 cohort, 0.343 to 2.577 in the ADNI cohort, and 0.428 to 3.063 in the meta-analysis. About half of the significant CpGs (21 CpGs) showed hypermethylation associated with an increased risk of dementia. Around half of these CpGs (20 CpGs) are located in CpG islands or shores. Additionally, 19 CpGs (43.18%) are in promoter regions within 2 kb of the TSS, which is significantly higher than the overall proportion of CpGs in promoter regions (27.65%) (*P-*value = 0.0276; Supplementary Figure 6).

**Figure 1.**
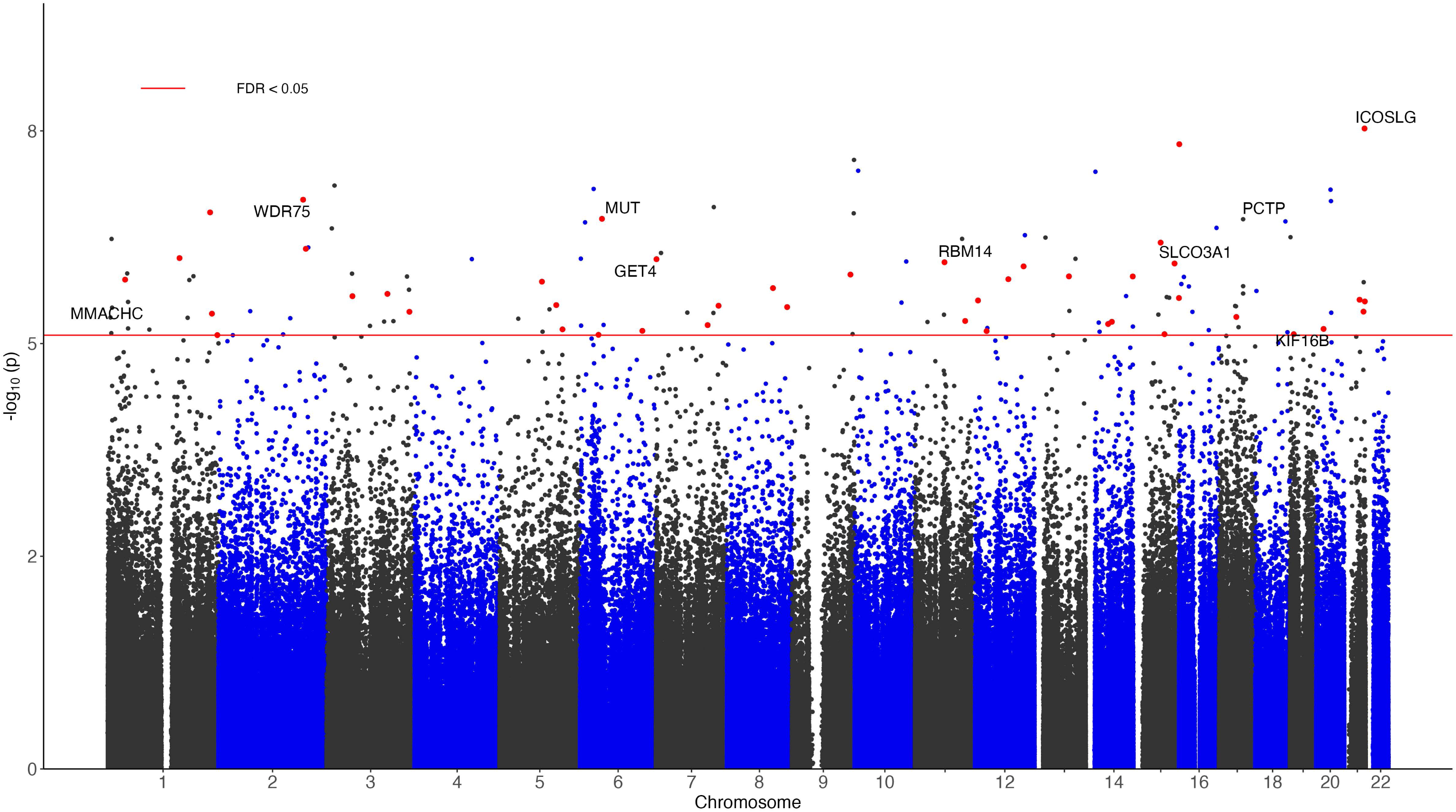
Manhattan plot of significant DNA methylation differences associated with dementia in meta-analysis of FHS9 (FHS at exam 9) and ADNI datasets. The X-axis indicates chromosome number. The Y-axis shows –log_10_(*P-*value) of meta-analysis, with red line indicating a 5% False Discovery Rate (FDR). The genes with promoter regions containing the top 10 most significant CpGs are highlighted. The red dots correspond to the 44 CpGs with a consistent direction of change in both FHS9 and ADNI datasets, a nominal *P*-value less than 0.05 in both datasets, and an FDR < 0.05 in meta-analysis of the FHS9 and ADNI datasets.

Using meta-analysis *P-*values for individual CpGs as input, comb-p^35^ software identified 44 differentially methylated regions (DMRs), which had a nominal *P*-value < 1×10^-5^, Sidak adjusted *P-*value < 0.05, and all the CpGs within the DMR have a consistent direction of change in estimated effect sizes in the meta-analysis (Supplementary Table 4). The number of CpGs in these DMRs ranged from 3 to 12. Among these DMRs, the majority showed hypermethylation associated with increased risk of dementia (35 DMRs), are located in CpG islands or shore (23 DMRs), or the promoter region (31 DMRs). Interestingly, among the significant individual CpGs and DMRs, 10 CpGs and 15 DMRs were also located in enhancer regions (Supplementary Table 3,4), which are regulatory DNA sequences that transcription factors bind to activate gene expression^38,48^

### Pathway analysis revealed DNA methylation differences associated with risk of dementia are enriched in biological pathways involved in immune responses and metabolic processes

To better understand biological pathways enriched with significant DNA methylation differences, we next performed pathway analysis using the methylGSA software^39^. At 5% false discovery rate (FDR), we identified 28 KEGG pathways and 26 Reactome pathways (Figure 2, Supplementary Table 5). Notably, a number of these significant pathways highlighted the central role of neuroinflammation processes in dementia, such as *B cell receptor signaling*, *Chemokine Signaling, Leukocyte Transendothelial Migration, Interleukin-1 Signaling*, and *Toll-like Receptor Cascades* pathways. In addition, several other significant pathways are involved in metabolic processes, including glucose and lipid metabolism, dysfunction of which are major risk factors for dementia ^49,50^. These significant pathways included *Glycolysis / Gluconeogenesis*, *Insulin signaling*, and *Steroid biosynthesis*.

**Figure 2.**
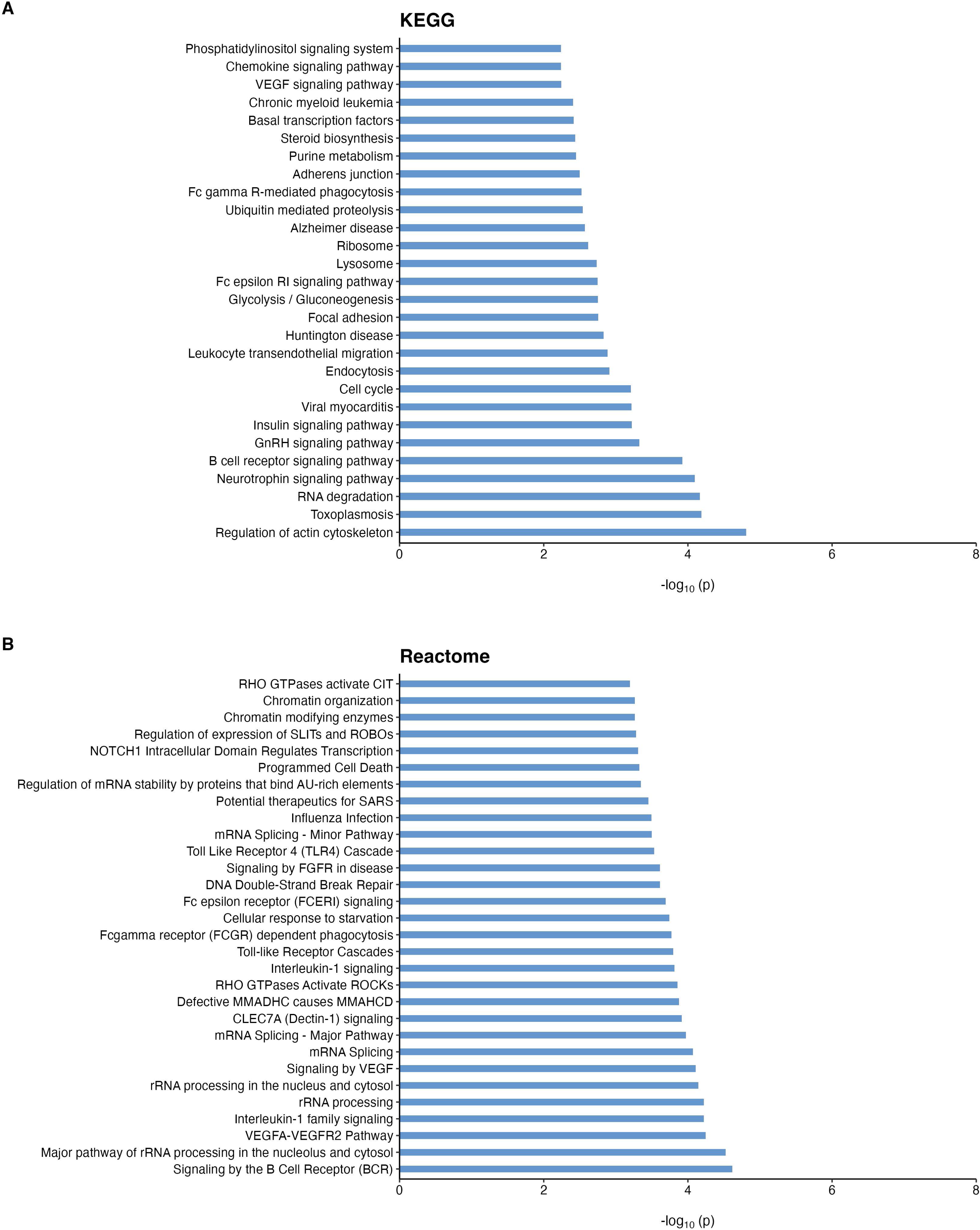
Significant KEGG (A) and Reactome (B) pathways enriched with dementia-associated CpGs at false discovery rate (FDR) less than 0.05.

### Correlation of significant DNAm with expression of nearby genes and brain DNA methylation levels

To better understand the functional role of the significant DMRs and CpGs, we performed several comparative analyses. We first overlapped our significant DNAm differences with previously established DNAm to RNA associations (i.e., eQTm) which was identified using matched DNA methylation and gene expression data from the Framingham study ^41^. Among the 44 significant individual CpGs and those within the 44 DMRs, we found 13 and 15 CpGs significantly correlated with target gene expression in *cis* (i.e., within 500k bp of the CpG) or *trans*, respectively (Supplementary Table 6). Notably, all the CpGs with *cis* associations are negatively correlated with their target gene expressions. Among them, 6 CpGs in the promoter regions of the *KIF16B* gene are significantly associated with its gene expression.

As dementia is a brain disorder, we also sought to prioritize methylation differences with a consistent direction of change in both blood and brain. To this end, we computed Spearman rank correlations between DNA methylation levels in the brain and blood using the London dataset^9^, which included matched DNA methylation samples measured on postmortem brain and pre-mortem blood samples of 69 subjects ^44^. We performed both an adjusted correlation analysis based on methylation residuals (*r_resid_*_u_) which removed covariate effects (see details in Methods) and an unadjusted correlation analysis based on methylation beta values (*r_beta_*). Among the significant individual CpGs and CpGs mapped within the DMRs, only 8 CpGs showed significant brain-to-blood associations in both adjusted and unadjusted analyses (*FDR*_*beta*_ < 0.05, *FDR*_resid_ < 0.05) (Supplementary Table 7). All of these CpGs were located in DMRs, and 6 out 8 CpGs showed significant positive brain-to-blood correlations. Notably, the three CpGs with the most significant brain-to-blood correlations (*r*_*beta*_: 0.821 to 0.851; *FDR*_*beta*_ 3.52×10^-16^ to 6.02×10^-19^) are located on the *ZNF696* gene, which encodes a zinc finger protein involved in transcriptional regulation (Supplementary Figure 7).

### Correlation and overlap with genetic risk loci

We identified methylation quantitative trait loci (mQTLs) by comparing our dementia-associated CpGs with blood mQTLs from the GoDMC database^42^. Among the 44 significant CpGs (Supplementary Table 3) and the 233 CpGs located in significant DMRs (Supplementary Table 4), 96 CpGs had 20974 mQTLs in *cis* and 12 CpGs had 1464 mQTLs in *trans* in the blood (Supplementary Table 8).

Next, we evaluated if the mQTLs overlapped with genetic risk loci implicated in dementia, by comparing them with the genetic variants nominated in a recent ADRD meta-analysis ^43^. We found that while no mQTLs overlapped with the genome-wide significant loci, 272 SNPs overlapped with genetic variants reaching a suggestive genome-wide significance threshold at *P* < 10^-5^ (Supplementary Table 9).

Given the observed overlap between the mQTLs and ADRD genetic risk loci, we next sought to determine whether the association signals at these loci (variant to CpG methylation levels and variant to clinical ADRD status) were due to a single shared causal variant or distinct causal variants close to each other. To this end, we performed a co-localization analysis using the method described by Giambartolomei et al. (2014)^51^. The results of this co-localization analysis strongly suggested^52^ (i.e. PP3+PP4 > 0.90, PP4 > 0.8 and PP4/PP3 > 5) that 9 genomic regions included a single causal variant common to both phenotypes (i.e. ADRD status and CpG methylation levels). These causal variants are located in the *IL34*, *CCR5AS* genes and the HLA intergenic regions (Supplementary Table 10).

### Sensitivity analyses

A growing body of recent research suggests that various lifestyle factors, such as smoking, may contribute to dementia ^53,54^. Meanwhile, recent studies also reported that DNA methylation is influenced by these lifestyle risk factors ^55–60^. We investigated whether any of the significant CpGs were also associated with dementia risk factors collected by the Framingham study, including *APOE*, diabetes, hypertension, years of education, BMI, and smoking. We found that among the 271 dementia-associated CpGs (44 significant individual CpGs, 233 CpGs located in significant DMRs, and 6 overlapping CpGs), 43 CpGs are associated with number of *APOE4* alleles, 28 CpGs are associated with smoking status, and 6 CpGs are associated with years of education. Moreover, 13, 18, and 7 CpGs are associated with BMI, diabetes, and hypertension status, respectively (Supplementary Table 11).

To evaluate the confounding effects of these risk factors, we re-analyzed the FHS9 dataset and performed a sensitivity analysis of the dementia-associated CpGs, by additionally adjusting for the significant risk factors they were associated with in the Cox regression model. Supplementary Table 12 shows the estimated hazard ratios (HRs) for all dementia-associated CpGs based on the original model and expanded model are very similar. Moreover, the *P-*values for the significant individual CpGs ranged from 1.20×10^-7^ to 4.02×10^-3^ in the original model and ranged from 2.20×10^-7^ to 0.0135 in the expanded model, indicating these CpGs are associated with dementia independent of the covariate factors. Given the importance of smoking as a confounding factor for DNAm studies, we also repeated the meta-analysis by additionally including smoking history as a covariate variable. Supplementary Table 13 shows that all 44 dementia-associated CpGs from Supplementary Table 3 remained highly significant, with meta-analysis *P-* values ranging from 1.79 ×10^-8^ to 2.15 ×10^-5^, corroborating the above results.

A second sensitivity analysis was performed to evaluate the impact of family structure in the FHS9 dataset on our analysis results. To this end, we estimated the intraclass correlation coefficient (ICC) for the dementia-associated CpGs, by comparing between-family variance to the total variance, which is the sum of between-family variance and within-family variance. Our results showed that for the 271 dementia-associated CpGs, the ICC values ranged from 0 to 0.149 (Supplementary Table 14), indicating minimal intraclass correlation in DNA methylation at these CpGs due to family structure.

In addition, we also performed an additional analysis using mixed-effects Cox models that accounted for family relationships with a kinship matrix computed from pedigree information. Notably, the *P-*values for the 44 dementia-associated individual CpGs ranged from 2.29×10^-9^ to 6.02×10^-3^ in the original model, and from 7.51×10^-8^ to 6.02×10^-3^ in the mixed effects Cox model. Supplementary Table 15 shows the hazard ratios and *P-*values from the mixed effects Cox model are very similar to those from the original Cox model, indicating that our results are robust to family structure in the FHS9 dataset.

In a third sensitivity analysis, we excluded CN subjects with strong biomarker evidence for AD, but short follow-up durations. These individuals are at a higher risk of progressing to dementia but may have incomplete data due to the limited follow-up period, which could introduce bias into the analysis. The FHS9 dataset includes plasma total tau measurements, while the ADNI dataset provides CSF pTau181 levels. Given the lack of consensus on specific cutoff values for AD biomarkers, we excluded subjects with tau biomarker levels in the highest quartile and follow-up times in the lowest quartile (i.e., ≤ 3 years). Supplementary Table 16 shows that the meta-analysis *P-*values for all 44 CpGs remained highly significant, ranging from 4.41×10^-8^ to 1.14×10^-5^, after excluding these high-risk subjects.

To evaluate the robustness of our DMR analysis results, we performed an additional DMR analysis on the genomic regions defined by the 44 significant DMRs listed in Supplementary Table 4, using an alternative approach, the coMethDMR software ^46^. Specifically, within each of these 44 genomic regions, we first identified co-methylated and differentially methylated regions associated with incident dementia, adjusting for age, sex, and blood cell-type composition for each dataset separately. We then combined the dataset-specific *P-*values for each genomic region using an inverse-variance fixed effects meta-analysis model. Our findings indicated that, among the 44 significant DMRs identified by comb-p, the majority (36 DMRs, 81.8%) were significantly associated with incident dementia using coMethDMR at a 5% FDR (Supplementary Table 17). Additionally, all 36 corroborated DMRs showed the same direction of effect in the coMethDMR method as in the comb-p method.

### Validation of dementia-associated CpGs in independent datasets

To validate our findings, we compared our dementia-associated CpGs and DMRs with those identified in previous studies using our recently developed MIAMI-AD database^47^. Our comparison revealed that 17 of the 44 significant individual CpGs (38.6%) overlapped with significant findings in previous research, with consistent direction of change (Supplementary Table 18). Among them, results for 9 CpGs were from independent cohorts other than FHS and ADNI. These 9 CpGs are located in the promoter regions of the *SLCO3A1, MUT, WDR75* genes and intergenic regions. Similarly, among the 227 CpGs located in DMRs, 65 CpGs were supported by previous research, also showing the same direction of effect. Among them, 51 CpGs located in 18 DMRs, reached nominal significance with the expected direction of change in external cohorts other than ADNI and FHS. These 18 DMRs were located on in the promoter regions of the *ACY3, ARMC5, CCR5, GET4, GLRX, IRAX4, KIF16B, LRRC59, C6orf25, MMACHC, HES5, PCTP, VAV1, ZMAT2*, *ZNF696* genes, and intergenic regions.

### Out-of-sample validation demonstrated the Methylation Risk Scores predicted dementia progression

To evaluate the feasibility of using dementia-associated DNAm for predicting disease progression, we developed an MRS score based on significant individual CpGs from the FHS9 dataset and assessed its ability to predict dementia progression in the ADNI dataset.

First, using the FHS9 dataset, we fitted a ridge regression model with time to dementia as the outcome and methylation *M-*values of 151 CpGs that achieved *P-*value < 10^-5^ in FHS9 dataset (using the model described above in Methods under “Association of DNA methylation at individual CpGs with dementia”) as predictors. Ridge regression reduces model variance by imposing a penalty on the size of the coefficients, leading to more stable and generalizable predictions. The estimated coefficients (i.e., weights) from ridge regression for the 151 CpGs ranged from −0.894 to 0.997, with positive weights assigned to 105 CpGs and negative weights to 46 CpGs (Supplementary Table 19). Notably, for all 151 CpGs, the directions of these weights were consistent with the estimated effect sizes from univariate Cox regression models that included each CpG individually. Moreover, the weights from ridge regression were significantly correlated with the estimated effect sizes from the univariate Cox regression models (Spearman correlation = 0.653, *P-*value < 2.2 ×10^-16^), supporting the robustness of the directionality of DNAm at these CpGs.

We next performed an out-of-sample validation of the MRS, a weighted sum of methylation *M-*values of the 151 CpGs, using an external dataset from the ADNI study, which included 538 subjects with available DNAm data and follow-up visit information (Supplementary Table 20). For each subject, we analyzed the earliest available (baseline) DNAm sample and additionally required the subjects to be CN or MCI at that time. These subjects were followed for an average of 5.39 ± 2.94 years, with follow-up durations ranging from 0.44 to 11.71 years. By their last visit, 64 (30.0%) CN subjects had progressed to MCI or Dementia, while 131 (40.3%) of the MCI subjects had progressed to dementia. Notably, the majority of these subjects (182 out of 195 subjects, 93.3%) who progressed to MCI or dementia did so due to AD.

For each of these 538 subjects, we computed MRS scores using DNAm data from their baseline visit and evaluated their association with disease progression (i.e., CN to MCI/dementia, MCI to dementia) using the Cox regression model. The MRS was computed by summing the methylation *M-*values of the 151 CpGs weighted by their coefficients estimated from FHS9 using ridge regression model. Table 4 shows that after adjusting for age, sex, *APOE ε4* status, years of education, baseline diagnosis, and baseline MMSE score, the MRS was significantly associated with progression to the next disease stage (estimate = 0.142, *P-*value = 0.041). Figure 3 shows survival probabilities over time for subjects in the highest and lowest MRS quartiles. Notably, while survival probability decreases in both groups, the group with low MRS consistently shows a higher survival probability, indicating lower risk of dementia progression. Similarly, when the analysis was limited to dementia due to AD using the same model, the MRS remained significant (estimate = 0.165, *P-*value = 0.0206) (Supplementary Table 21) and the survival probabilities showed similar trends (Supplementary Figure 8).

**Figure 3.**
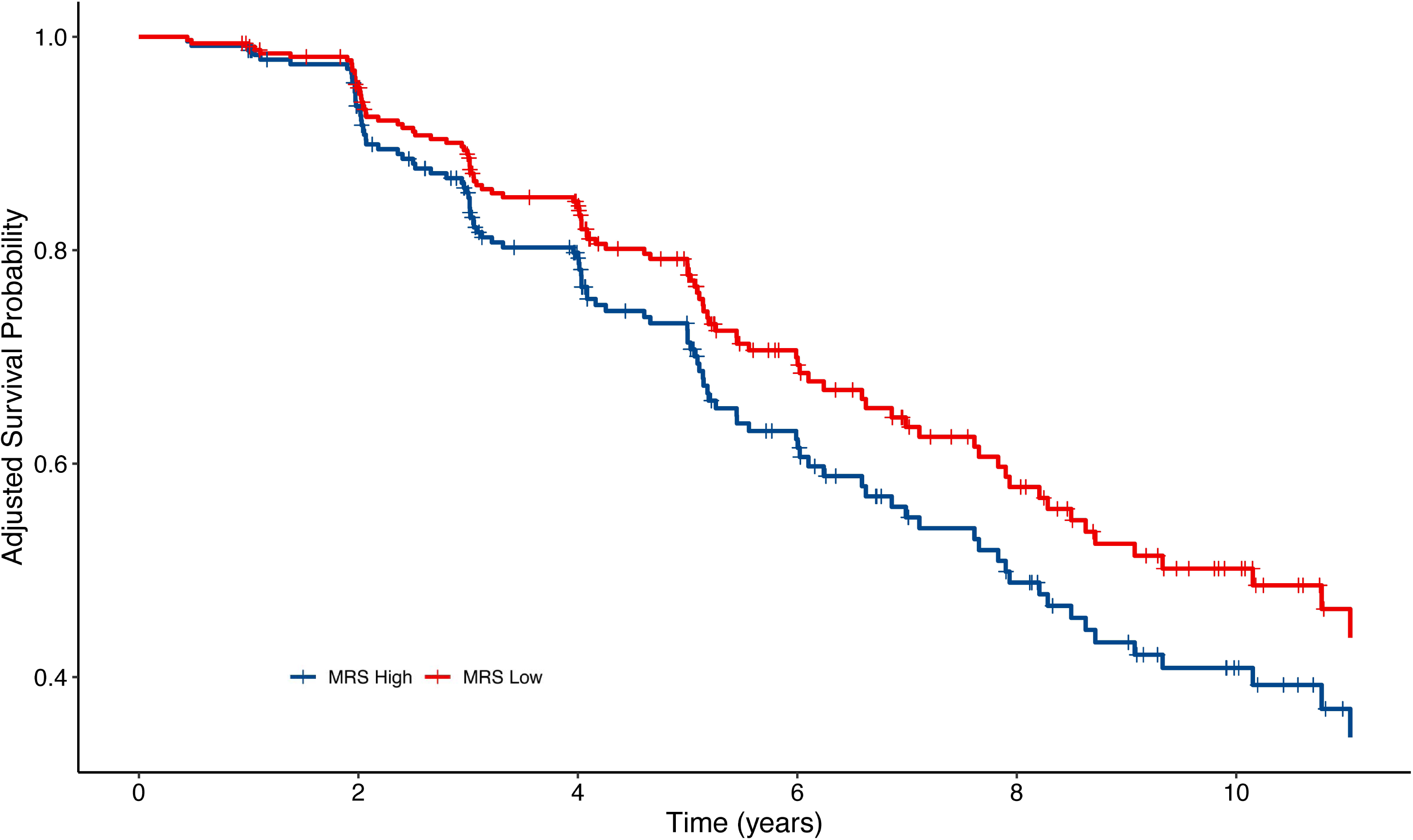
Adjusted Kaplan-Meier curves for dementia progression (CN to MCI/dementia, or MCI to dementia) among subjects in the highest and lowest quartiles of baseline MRS scores in the ADNI cohort. The survival probability decreases in both groups over time. Moreover, the group in the lowest MRS quartile consistently shows a higher survival probability, indicating a lower risk for dementia progression. These results are adjusted for age, sex, APOE ε4, years of education, baseline diagnosis, and baseline MMSE score. **Abbreviations** CN, cognitively normal; MCI, mild cognitive impairment; MRS, Methylation Risk Score

**Table 4.** Results from Cox regression model evaluating the association between Methylation Risk Score (MRS) and disease progression (CN to MCI/dementia, MCI to dementia) in 538 subjects, adjusted for age, sex, APOE ε4 status, baseline diagnosis, MMSE, and education using ADNI dataset. Significant association was observed for MRS (estimate = 0.142, *P-*value = 0.0413), indicating higher MRS increases risk.

| Characteristic | Coefficient | HR (95% CI) | P-value |
| --- | --- | --- | --- |
| **MRS** | **0.142** | **1.152 (1.006,1.32)** | **0.041** |
| **Age, years** | **0.066** | **1.068 (1.047,1.091)** | **3.76 × 10<sup>-10</sup>** |
| **Male** | **0.062** | **1.064 (0.786,1.44)** | **0.690** |
| **Baseline diagnosis** |  |  |  |
| CN |  | 1 [Reference] |  |
| MCI | **0.494** | **1.639 (1.177,2.281)** | **3.44 × 10<sup>-3</sup>** |
| **APOE ε4 allele** | **0.646** | **1.907 (1.531,2.376)** | **8.58 × 10<sup>-9</sup>** |
| **MMSE** | **-0.180** | **0.835 (0.762,0.916)** | **1.26 × 10<sup>-4</sup>** |
| **Education, years** | **-0.002** | **0.998 (0.942,1.058)** | **0.949** |

**Abbreviations:** MRS, Methylation Risk Score; HR, hazard ratio; CI, confidence interval; CN, cognitively normal; MCI, Mild Cognitive Impairment; MMSE, Mini-Mental State Examination

## DISCUSSION

We performed a comprehensive analysis of more than 1000 blood samples to identify DNA methylation associated with incident dementia in two longitudinal studies. After correcting for multiple comparisons, we identified 44 CpGs and 44 DMRs significantly associated with dementia risk (Supplementary Tables 3-4). Comparing these significant DNAm differences with findings from previous cross-sectional studies, we found that approximately 40% of the significant CpGs and 30% of the DMRs overlapped with previous results. While differences in sample characteristics, the specific arrays used, and the statistical models applied may contribute to this discrepancy, it also suggests that some DNAm differences observed in cross-sectional studies could be due to reverse causation of the disease. The novel DNAm differences discovered in this study highlighted the importance of using a longitudinal design to identify DNAm changes with a temporal relationship to the disease.

Importantly, a number of our findings pointed to early processes in dementia, where DNA methylation could serve as biomarkers and potential therapeutic targets can be developed. For example, our results confirmed the central role of neuroinflammation in dementia^61–63^, including immune responses to dementia pathology such as amyloid beta, which may be deposited in the brain decades before the onset of clinical symptoms ^64^. Our most significant CpG is located on the *ICOSLG* gene associated with T-cell activation ^65–67^. Additionally, two of the top 10 most significant DMRs are located on the *IRAK4* and *ACY3* genes, which are associated with microglia activation ^68,69^. Our pathway analysis also pointed to immune responses to dementia pathology, highlighting pathways such as *B cell receptor signaling*, *Chemokine Signaling, Leukocyte Transendothelial Migration, Interleukin-1 Signaling*, and *Toll-like Receptor Cascades*. It has been proposed that reducing neuroinflammation may be a promising strategy for delaying the onset and progression of neurodegenerative diseases ^70,71^.

Our results also underscore the role of metabolic dysfunction as an early event in dementia, well before significant amyloid-beta protein accumulation^72,73^, which is consistent with late-onset diabetes being an established risk factor for dementia ^74^. Our most significant DNAm included CpGs and DMRs located on the *MUT*, *GET4*, and *NUDT19* genes which are crucial for mitochondrial function and energy metabolism. The most significant DMR is located in the *PCTP* gene involved in lipid metabolism, which is increasingly recognized for its important role in Alzheimer’s dementia^75^. Our pathway analysis also highlighted a number of metabolic processes, including *glycolysis/gluconeogenesis*, *insulin signaling*, and *steroid biosynthesis*. Impairments in these pathways lead to energy deficits, oxidative stress, synaptic dysfunction, and inflammation, which are early hallmarks of neurodegeneration. Encouragingly, recent research suggests that targeting mitochondria may offer promising therapeutic targets for the treatment and prevention of dementia ^76^. In model organisms, the removal of defective mitochondria diminishes insoluble Aβ1-42 and reverses memory impairment^77^, and treatment with anti-diabetes drugs reduces protein aggregation and reverses Aβ-induced metabolic defects ^78^. Moreover, insulin treatment improved cognitive function in subjects with mild cognitive impairment ^79^.

To better understand the genetic influences on dementia-associated DNAm, we leveraged the GoDMC database, which includes blood mQTLs computed from 32,851 independent subjects^42^. Consistent with previous observations that genetic influences on DNAm in the blood are widespread ^80^, we found that about 40% (99 out of 271 CpGs) of our dementia-associated CpGs are associated with mQTLs. This is similar to findings by Min et al. (2021), who estimated that genetic variants influence about 45% of DNAm sites on the Illumina array^42^. To prioritize potential regulatory variants causally involved in dementia, we performed a co-localization analysis of association signals from two independent studies: the mQTLs of dementia-associated DNAm identified in this study and genetic risk loci implicated in a recent ADRD GWAS ^43^. The co-localization analysis revealed that a subset of CpGs are influenced by mQTLs (i.e., SNPs) linked to ADRD risk. The significant co-localization signals at these loci provided strong support that ADRD and CpG methylation are associated with the same causal genetic variant. These nominated causal genetic variants are located on the *IL34, CCR5AS* genes and the HLA intergenic region, all of which are involved in neuroinflammatory responses relevant to dementia. Future studies using multi-omics data, including genetic variants, DNAm, and dementia outcomes, are needed to thoroughly investigate and confirm whether the genetic associations with AD at these loci are mediated by methylation modifications at nearby CpG sites. Alternatively, these DNAm differences might simply be peripheral markers of indirect systemic effects reflecting the underlying AD-related pathology. Given that the primary pathology of AD occurs in brain tissue, future studies of mQTLs and co-localization in both brain and blood samples, especially cell-type-specific studies, will provide additional insights into the genetic regulatory mechanisms by which the dementia-associated loci may influence DNAm to affect disease risk.

Compared to gene expression and proteins, methylated DNA is relatively stable and can be easily detected, thus serving as an excellent source of biomarkers^81^. Recently, a number of blood-based AD biomarkers, such as the high-performing plasma pTau217, have been developed and shown to be significantly associated with the future development of AD dementia in subjects with MCI ^82^. However, autopsy studies have observed a discordance between neuropathological burden and cognitive performance^83^. These studies have revealed that among CN subjects, about a quarter exhibit amyloid abnormalities in the brain that meet the neuropathological criteria for AD^84,85^. Similarly, current pathology biomarkers do not completely predict subsequent cognitive impairment in asymptomatic individuals. For example, in a recent study by Ossenkoppele et al. (2022), among CN subjects identified with both amyloid and tau pathology (A+T+), as measured by amyloid-PET and tau-PET, which indicates a high likelihood of progression to MCI or AD, a substantial proportion (∼36%, 40 out of 111) remained CN after an average of 42 months of follow-up ^86^.

Cognitive resilience (CR) refers to the adaptability of an individual to brain changes due to disease, injury, or normal aging ^87,88^. Higher CR has been associated with a lower risk of progression to clinical AD and a slower rate of cognitive decline^89^. Lifestyle factors such as engaging in physical activity and following the MIND diet, which have been shown to be associated with DNA methylation, play important roles in promoting CR and delaying cognitive decline ^90–93^. Therefore, DNA methylation holds great promise as a complementary biomarker to existing pathology biomarkers, facilitating a more precise determination of dementia risk.

The strengths of this study include the longitudinal design of the FHS and ADNI studies, which allowed us to identify DNAm associated with incident dementia. In both studies, dementia status was adjudicated by a team of experts based on comprehensive data, including clinical assessments, cognitive testing, and biomarkers. The DNAm samples in both ADNI and FHS9 studies were measured using Illumina EPIC arrays, which provide improved coverage of regulatory elements ^94^ and were recently shown to generate more reliable DNA methylation levels than the older 450k arrays ^95^. To reduce concerns of false positives, we adjusted for potential confounding effects such as age, sex, estimated major immune cell-type proportions in the blood, and corrected for batch effects in our meta-analysis. We also performed inflation correction using the bacon method ^33^, specifically designed for epigenome-wide association studies. We also used stringent criteria to select our significant CpGs and DMRs. For significant individual CpGs, we required consistent directional effects and nominal significance in both cohorts. For DMRs, we required all CpGs within the DMR to have consistent directional effects. Moreover, we evaluated the sensitivity of our results to major risk factors of dementia, some of which also correlated with DNA methylation. We found that all of our 44 dementia-associated CpGs remained significant, indicating their association with dementia is independent of the risk factors. We also estimated intraclass correlation coefficients and performed additional analyses accounting for family relationships in FHS9 using a kinship matrix. The results of this analysis showed our findings are robust to family structure in the FHS9 dataset. Finally, and importantly, we showed that the Methylation Risk Score developed using the Framingham study dataset is significantly associated with progression to MCI or dementia in the ADNI dataset, even after accounting for age, sex, APOE ε4, years of education, baseline diagnosis, and baseline MMSE score. Recently, Koetsier et al. (2024) also developed a DNAm-based risk score that successfully predicted future cognitive impairment in independent AD and Parkinson’s disease datasets, by leveraging DNAm associated with 14 modifiable and non-modifiable dementia risk factors ^96^. Our study, which identified DNAm directly associated with incident dementia, and the study by Koetsier et al., which discovered DNAm associated with dementia risk factors, provided complementary approaches for discovering risk variants for dementia. Together, the results of our study and theirs demonstrate that DNAm is a plausible predictive biomarker that precedes dementia onset.

This study has several limitations. First, we analyzed bulk blood DNA methylation samples that contain a mixture of cell types. To reduce confounding effects due to cellular diversity, we included estimated cell-type proportions as covariates in all our analyses. Future studies using single-cell technology could provide more detailed insights into the specific cell types affected by the dementia-associated DNA methylation differences identified here. However, single-cell methylomic technologies currently face significant practical challenges. These methods typically involve high costs, complex data processing, and require substantial technical expertise, which can limit their scalability, especially for large, complex disease cohorts like those studied in ADRD research. Moreover, the heterogeneity inherent in ADRD complicates the application of single-cell approaches, as large sample sizes across diverse cell populations are needed to capture meaningful biological variability.

Second, an interesting observation is the distribution of hypermethylated CpGs and DMRs, where approximately half (21 out of 44) of the FDR-significant CpGs were hypermethylated, compared to three-quarters (35 out of 44) of the identified DMRs. This discrepancy may reflect potential biases inherent in array-based technology, which includes a higher density of probes within promoter regions, areas where DMRs are more likely to be detected. Such biases could have contributed to the observed higher proportion of hypermethylated DMRs. The use of sequencing-based technologies in future studies would help to overcome these biases by offering more comprehensive and unbiased coverage of the methylome.

Third, to help increase sample size, we used a broad definition of dementia to identify DNAm signatures, which might have diluted association signals due to the heterogeneity among various dementia subtypes. However, the majority of dementia events in our study were attributed to AD. In the FHS9 dataset, 32 of 42 (76.2 %) dementia events were due to AD, and in the ADNI dataset, 15 of 18 (83.3 %) dementia events were due to AD. When we repeated the meta-analysis of FHS9 and ADNI datasets using the same models but replacing time to incident dementia with time to AD dementia, the results were very similar. In particular, the meta-analysis *P-*value for all 44 CpGs in Supplementary Table 3 remained highly significant, ranging from 5.26 ×10^-8^ to 6.85×10^-3^ (Supplementary Table 22). Moreover, the MRS score developed using the FHS9 dataset, with time to dementia as the outcome variable, validated well in the ADNI dataset, and was significantly associated with progression to MCI or dementia due to AD, even after adjusting for covariate variables (Supplementary Table 21, Supplementary Figure 8).

Also, due to the lack of data, we analyzed only DNA methylation samples from non-Hispanic white subjects, and the subjects in both cohorts were highly educated. Future studies that investigate DNA methylation in large, multi-ethnic cohorts with diverse backgrounds are needed. Finally, while both ADNI and FHS have dementia surveillance programs, the insidious onset of dementia might lead to underreporting, with some subjects reaching dementia status before their recorded onset date. These cases could dilute the association signals between DNA methylation and incident dementia in our study, making our meta-analysis results conservative. It has been estimated that a substantial proportion of dementia cases may be undiagnosed or not reported ^2^. Therefore, a sensitive and objective biomarker, such as DNA methylation that can be easily quantified, is urgently needed to help improve surveillance of incident dementia.

In summary, we identified numerous DNA methylation differences consistently associated with incident dementia in a meta-analysis of two longitudinal cohorts, comprising over 1,000 blood samples. Our comparative analysis, which incorporated results from integrative analysis of blood DNA methylation with gene expression, genetic variants, and brain DNA methylation data, and pathway enrichment analysis highlights the central role of neuroinflammation and early processes such as metabolic dysfunction in dementia. Importantly, our out-of-sample validation demonstrated that methylation risk scores based on dementia-associated DNAm predicted future cognitive decline in an independent dataset, even after accounting for covariate variables, supporting blood DNAm as a potential objective biomarker for identifying individuals at higher risk for dementia. Future studies that validate our findings in larger and more diverse community-based cohorts are warranted.

## Data Availability

The genome-wide summary statistics have been deposited to the MIAMI-AD (DNA Methylation in Aging and Methylation in AD) database (https://miami-ad.org/)

https://github.com/TransBioInfoLab/blood-dnam-and-incident-dementia

## ACKNOWLEDGMENTS

We would like to thank Dr. Rhoda Au and her team for their valuable assistance in helping us to better understand the Framingham Heart Study dataset. Data collection and sharing for the ADNI dataset was funded by the Alzheimer’s Disease Neuroimaging Initiative (ADNI) (National Institutes of Health Grant U01 AG024904) and DOD ADNI (Department of Defense award number W81XWH-12-2-0012). ADNI is funded by the National Institute on Aging, the National Institute of Biomedical Imaging and Bioengineering, and through generous contributions from the following: AbbVie, Alzheimer’s Association; Alzheimer’s Drug Discovery Foundation; Araclon Biotech; BioClinica, Inc.; Biogen; Bristol-Myers Squibb Company; CereSpir, Inc.; Cogstate; Eisai Inc.; Elan Pharmaceuticals, Inc.; Eli Lilly and Company; EuroImmun; F. Hoffmann-La Roche Ltd and its affiliated company Genentech, Inc.; Fujirebio; GE Healthcare; IXICO Ltd.; Janssen Alzheimer Immunotherapy Research & Development, LLC.; Johnson & Johnson Pharmaceutical Research & Development LLC.; Lumosity; Lundbeck; Merck & Co., Inc.; Meso Scale Diagnostics, LLC.; NeuroRx Research; Neurotrack Technologies; Novartis Pharmaceuticals Corporation; Pfizer Inc.; Piramal Imaging; Servier; Takeda Pharmaceutical Company; and Transition Therapeutics. The Canadian Institutes of Health Research is providing funds to support ADNI clinical sites in Canada. Private sector contributions are facilitated by the Foundation for the National Institutes of Health (www.fnih.org). The grantee organization is the Northern California Institute for Research and Education, and the study is coordinated by the Alzheimer’s Therapeutic Research Institute at the University of Southern California. ADNI data are disseminated by the Laboratory for Neuro Imaging at the University of Southern California.

## CONSENT STATEMENT

The ADNI and Framingham Heart Study were approved by the institutional review boards of all participating institutions. Written informed consent was obtained from all the participants or their authorized representatives.

