## Supplementary Figures for "Blood DNA Methylation Signature for Incident Dementia: Evidence from Longitudinal Cohorts"

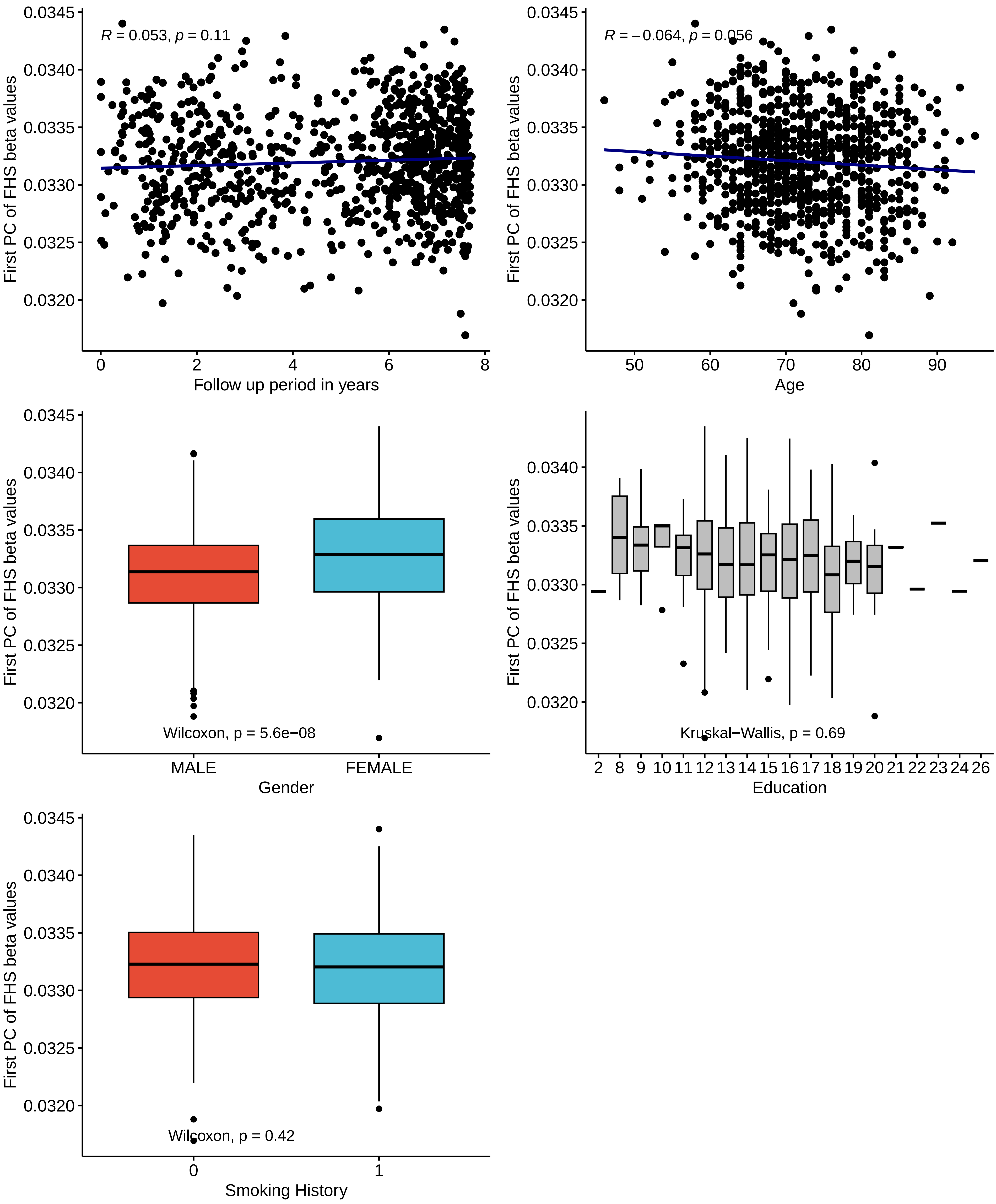


**Supplementary Figure 1** The first principal component (PC1) of DNA methylation beta values in the Framingham Heart Study Exam 9 dataset was not significantly associated with follow up duration, age, education, and smoking history. However, PC1 was significantly higher in males.

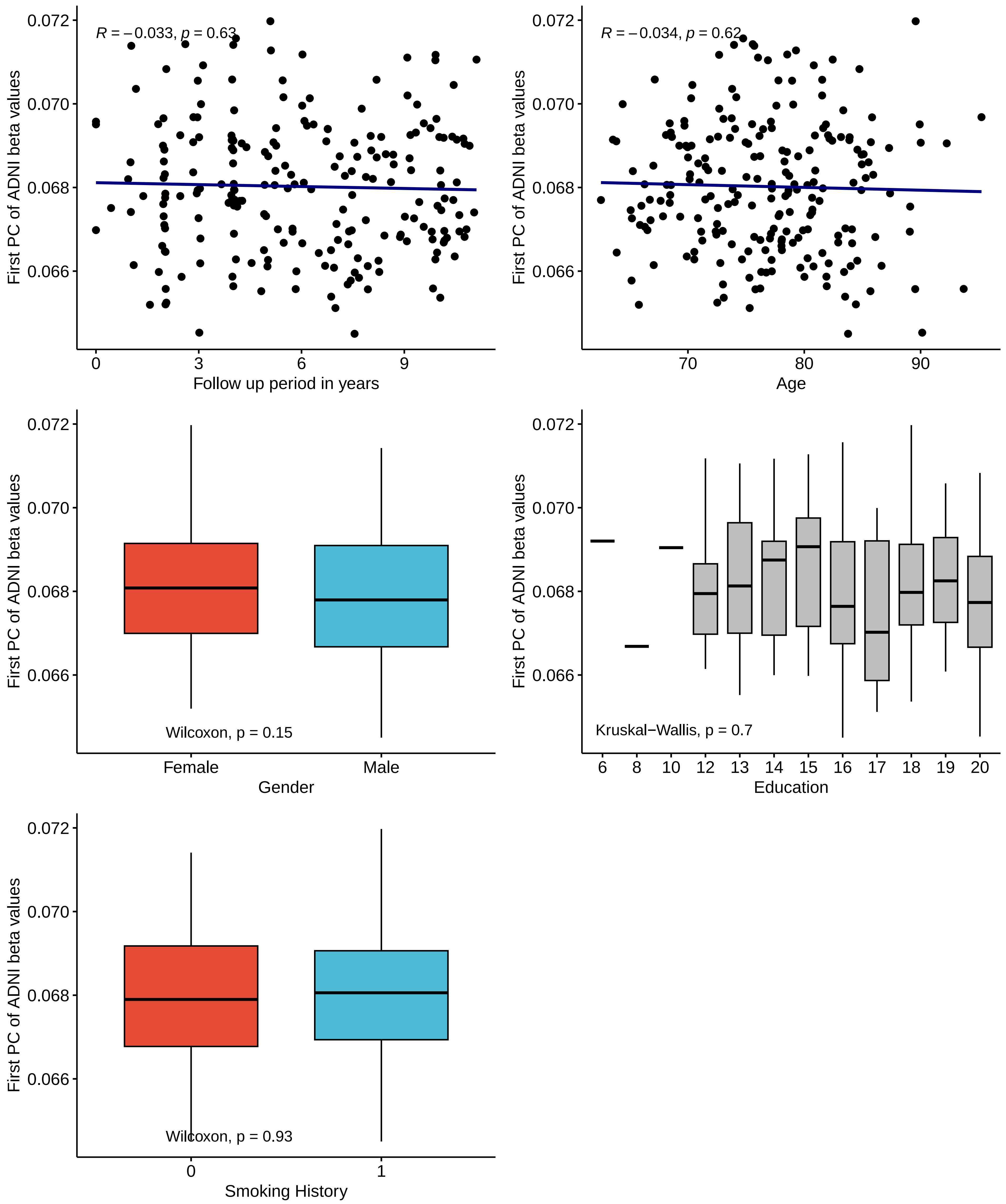


**Supplementary Figure 2** The first principal component of DNA methylation beta values in the ADNI dataset was not significantly associated with follow up duration, age, sex, education, and smoking history.

**Supplementary Figure 3** Histogram of Dementia Follow−Up Time in the Framingham Heart Study

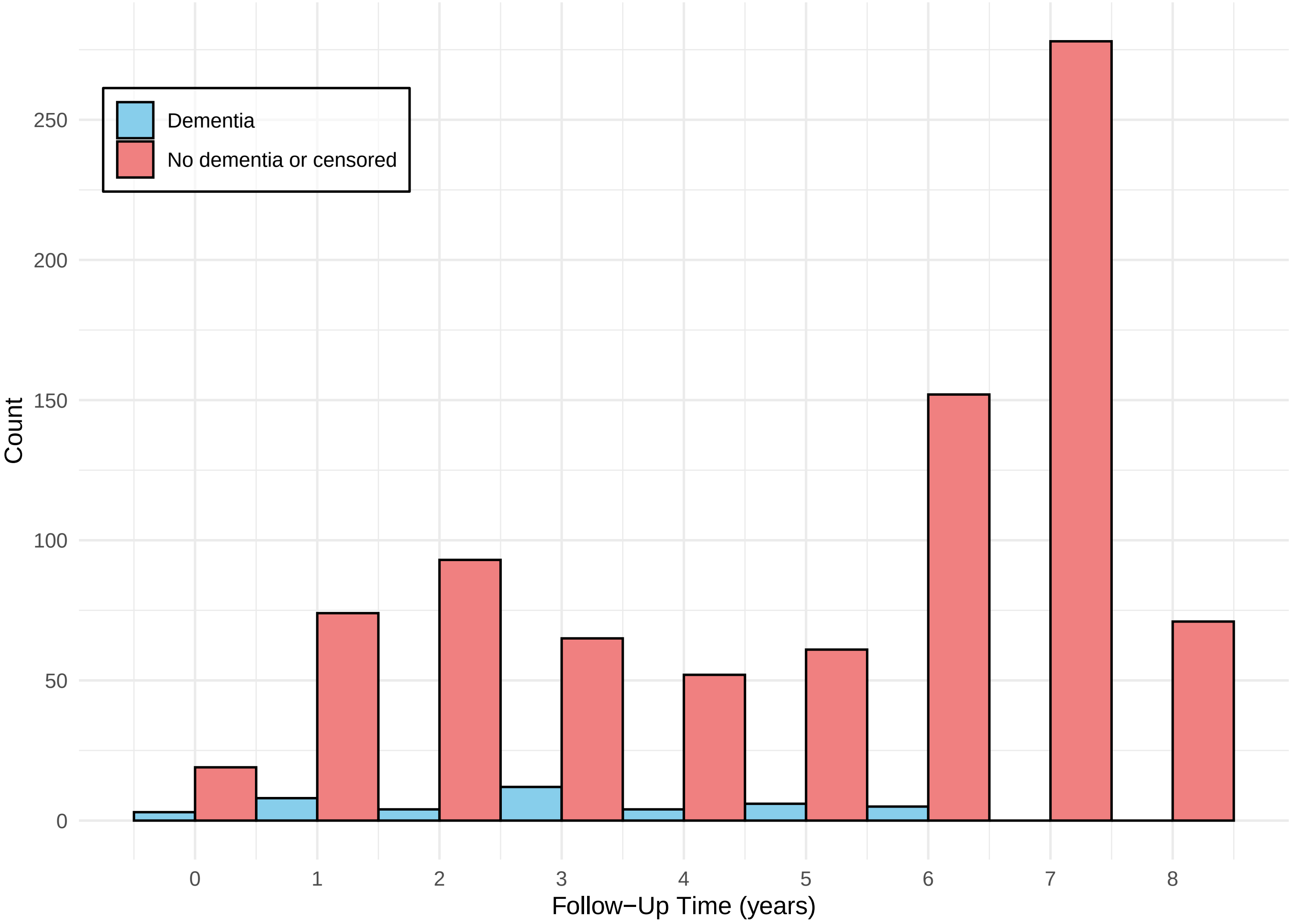

**Supplementary Figure 4** Histogram of Dementia Follow−Up Time in the ADNI Study

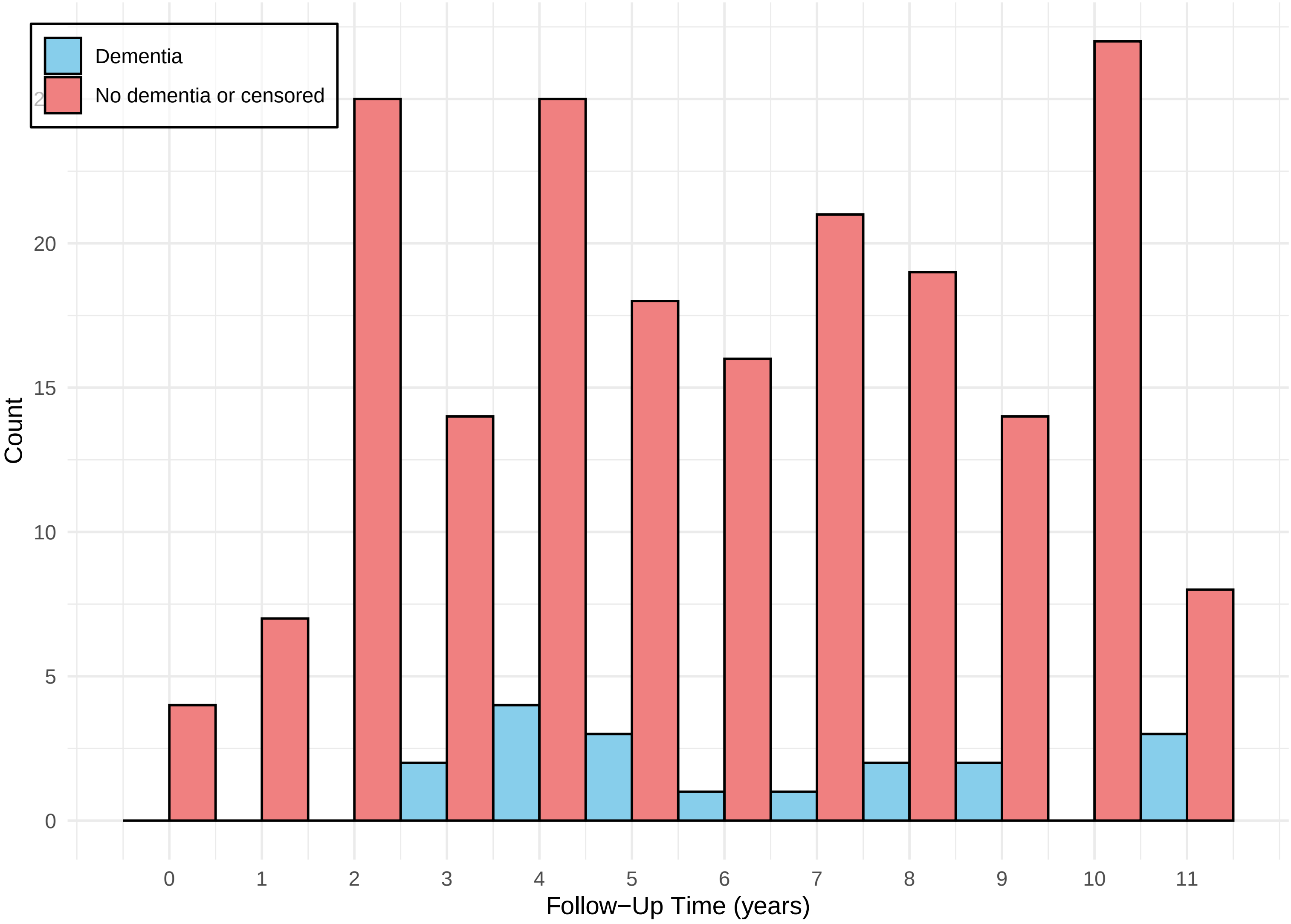

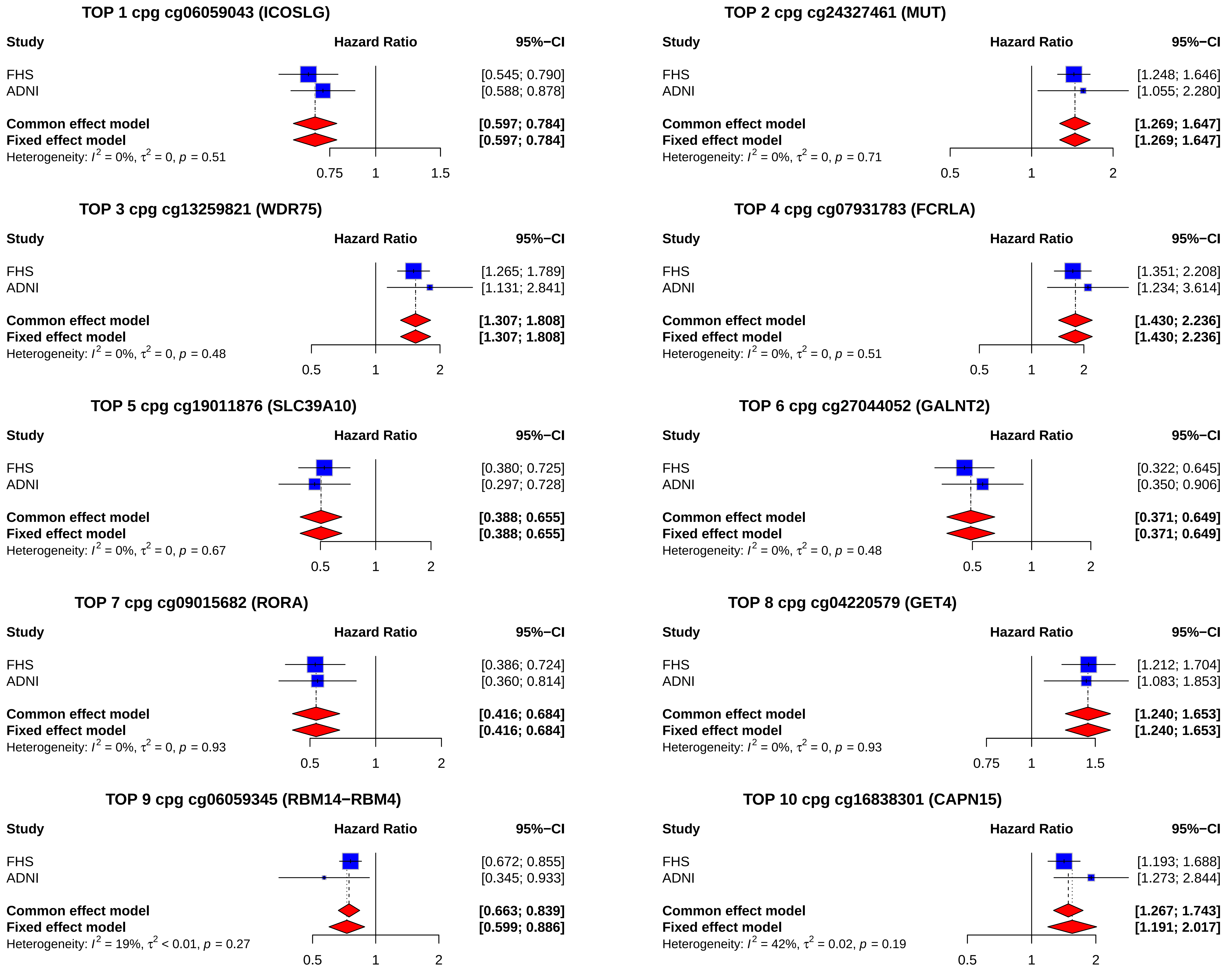


**Supplementary Figure 5** Forest plots for the top 10 most significant CpGs in meta-analysis of FHS9 (FHS at exam 9) and ADNI blood samples datasets. Shown are hazard ratios (HR) that describe changes in risk of dementia associated with one standard deviation increase in methylation beta values after adjusting for covariate variables (age, sex, and immune cell-type proportions).

**(A) Enrichment analysis of FDR-significant CpGs in meta-analysis of ADNI and FHS9 studies, with respect to CpG islands / shores and promoter regions** (i.e., less than ± 2k bp from TSS)

| Significant.CpG | CpG.island.or.shore Yes | No | Row Total |
|---|---|---|---|
| Yes | 20<br>45.455% | 24<br>54.545% | 44<br>0.006% |
| No | 285701<br>36.812% | 490401<br>63.188% | 776102<br>99.994% |
| Column Total | 285721 | 490425 | 776146 |

```
Fisher's Exact Test for Count Data
------------------------------------------------------------
Sample estimate odds ratio:  1.430352

Alternative hypothesis: true odds ratio is not equal to 1
p =  0.2737635
95% confidence interval:  0.7493822 2.702118
```

| Significant.CpG | Promoter.region Yes | No | Row Total |
|---|---|---|---|
| Yes | 19<br>43.182% | 25<br>56.818% | 44<br>0.006% |
| No | 214594<br>27.650% | 561508<br>72.350% | 776102<br>99.994% |
| Column Total | 214613 | 561533 | 776146 |

```
Fisher's Exact Test for Count Data
------------------------------------------------------------
Sample estimate odds ratio:  1.988704

Alternative hypothesis: true odds ratio is not equal to 1
p =  0.02757649
95% confidence interval:  1.035269 3.760547
```

**(B) CpGs located with respect to CpG islands and promoter regions**

| Relation to CpG island | Promoter region FALSE | TRUE | Row Total |
|---|---|---|---|
| Island | 48479<br>33.226% | 97429<br>66.774% | 145908<br>18.799% |
| N_Shelf | 25791<br>92.547% | 2077<br>7.453% | 27868<br>3.591% |
| N_Shore | 33861<br>44.908% | 41539<br>55.092% | 75400<br>9.715% |
| OpenSea | 401949<br>92.038% | 34774<br>7.962% | 436723<br>56.268% |
| S_Shelf | 24001<br>92.905% | 1833<br>7.095% | 25834<br>3.328% |
| S_Shore | 27452<br>42.619% | 36961<br>57.381% | 64413<br>8.299% |
| Column Total | 561533 | 214613 | 776146 |

**Supplementary Figure 6** The enrichment analyses showed the 44 FDR-significant CpGs were not significantly enriched in CpG islands or shores (*P*-value = 0.2738), but were significantly enriched in promoter regions (*P*-value = 0.0276) (A, B). Although the majority of CpGs located in islands or shores are also found in promoter regions, a substantial proportion of these CpGs are located outside promoter regions as well.

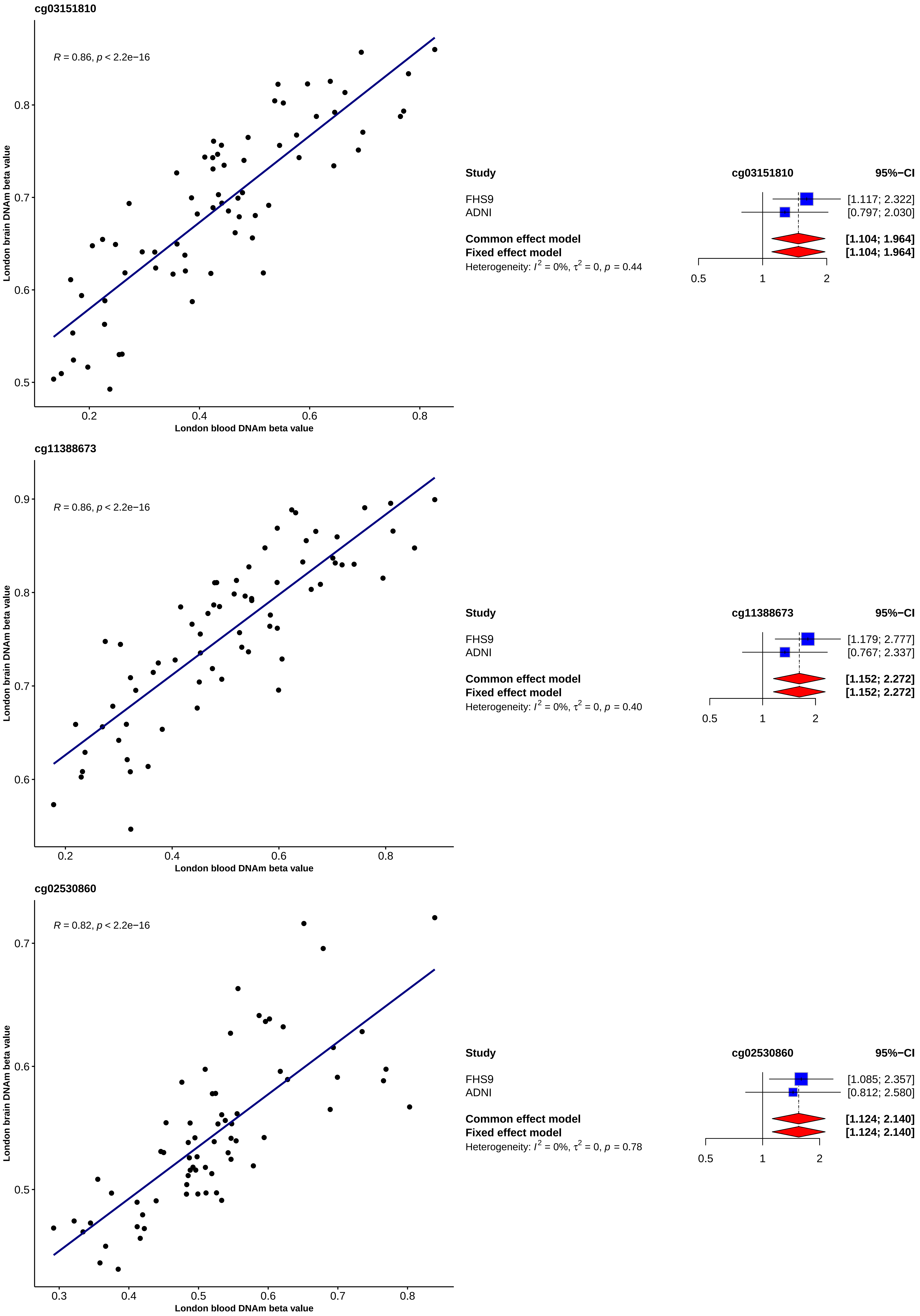


**Supplementary Figure 7** DNA methylation at three CpGs located in the promoter of the *ZNF696* gene showed significant brain-blood correlation in the London cohort samples (PMID: 26457534), and are are significantly associated with incident dementia in the meta-analysis of FHS9 (FHS at exam 9) and ADNI datasets.

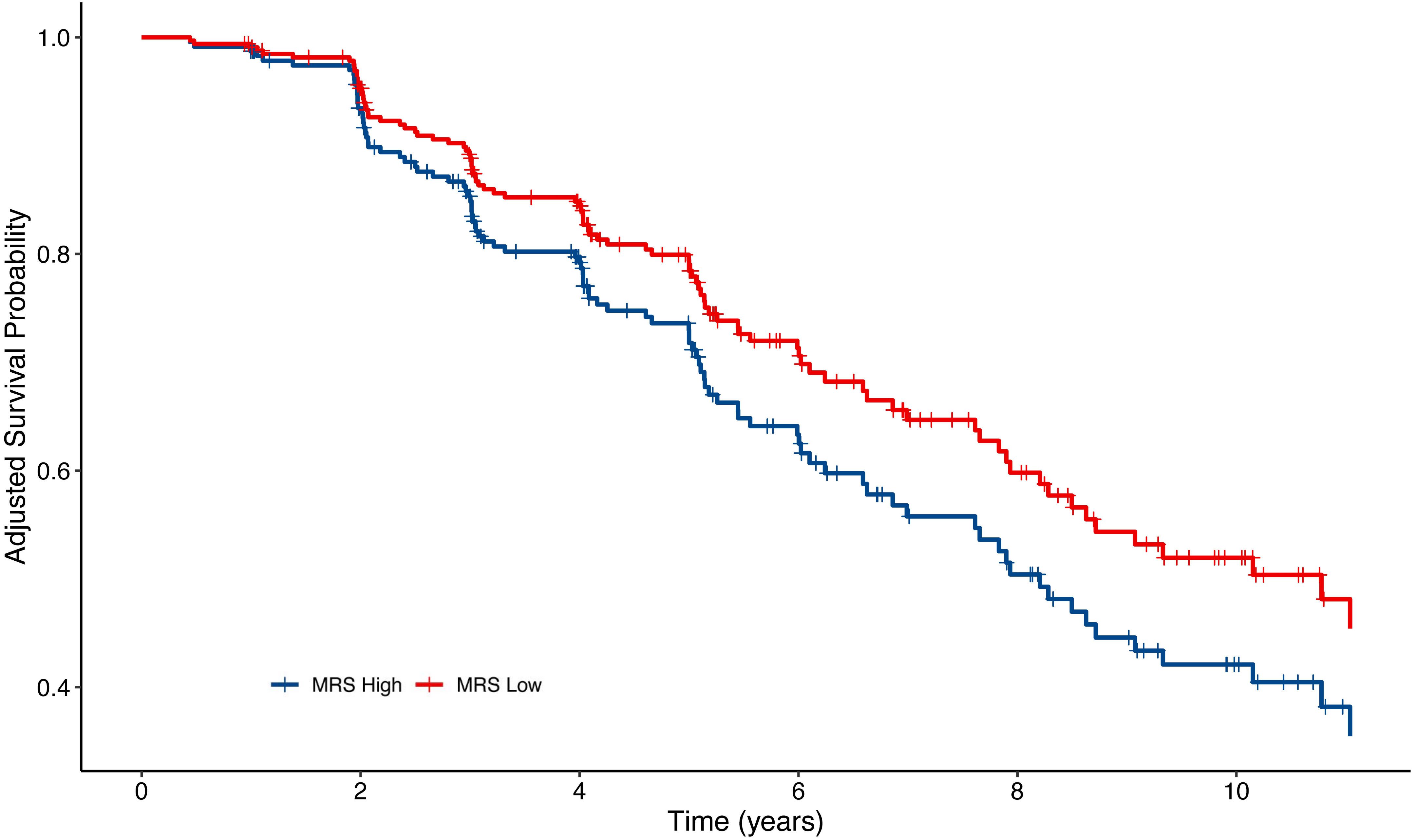


**Supplementary Figure 8** Adjusted Kaplan-Meier curves for AD dementia progression (CN to MCI/AD, or MCI to AD) among subjects in the highest and lowest quartiles of baseline MRS scores in the ADNI cohort. The survival probability decreases in both groups over time. Moreover, the group in the lowest MRS quartile consistently shows a higher survival probability, indicating a lower risk for dementia progression. These results are adjusted for age, sex, APOE ε4, years of education, baseline diagnosis, and baseline MMSE score.
**Abbreviations** CN, cognitively normal; MCI, mild cognitive impairment; AD, Alzheimer's disease; MRS, Methylation Risk Score
